# Evaluating AI Reasoning Models in Pediatric Medicine: A Comparative Analysis of o3-mini and o3-mini-high

**DOI:** 10.1101/2025.02.27.25323028

**Authors:** Gianluca Mondillo, Mariapia Masino, Simone Colosimo, Alessandra Perrotta, Vittoria Frattolillo

## Abstract

Artificial intelligence (AI) is increasingly playing a crucial role in modern medicine, particularly in clinical decision support. This study compares the performance of two OpenAI reasoning models, o3-mini and o3-mini-high, in answering 900 pediatric clinical questions derived from the MedQA-USMLE dataset. The evaluation focuses on accuracy, response time, and consistency to determine their effectiveness in pediatric diagnostic and therapeutic decision-making. The results indicate that o3-mini-high achieves a higher accuracy (90.55% vs. 88.3%) and faster response times (64.63 seconds vs. 71.63 seconds) compared to o3-mini. The chi-square test confirmed that these differences are statistically significant (X^2^ = 328.9675, *p <* 0.00001)). Error analysis revealed that o3-mini-high corrected more errors from o3-mini than vice versa, but both models shared 61 common errors, suggesting intrinsic limitations in training data or model architecture. Additionally, accessibility differences between the models were considered. While DeepSeek-R1, evaluated in a previous study, offers unrestricted free access, OpenAI’s o3 models have message limitations, potentially influencing their suitability in resource-constrained environments. Future improvements should aim at reducing shared errors, optimizing o3-mini’s accuracy while maintaining efficiency, and refining o3-mini-high for enhanced performance. Implementing an ensemble approach that leverages both models’ strengths could provide a more robust AI-driven clinical decision support system, particularly in time-sensitive pediatric scenarios such as emergency care and neonatal intensive care units.

## Introduction

Artificial intelligence (AI) is playing an increasingly central role in modern medicine, offering innovative solutions for clinical data management, diagnostic analysis, and therapeutic decision support. Reasoning models, in particular, represent a new frontier in medical AI, enabling the handling of complex problems through a logical and sequential process. [1] This study aims to compare the performance of two OpenAI reasoning models, o3-mini and o3-mini-high, on a set of 900 pediatric questions. The objective of the study is to systematically evaluate the accuracy, timeliness, and consistency of the responses provided by the analyzed models to identify which of them ensures the most effective clinical support, with particular attention to the pediatric context, given the complexity and significance of diagnostic and therapeutic decisions in this population. Reasoning models, such as o3-mini and o3-mini-high [2], differ from traditional language models due to their ability to segment complex problems into simpler logical steps, thus improving response reliability. DeepSeek-R1 [3], an open-source model based on reinforcement learning techniques, has already demonstrated high accuracy (87%) and good speed in our previous study [4], making it particularly suitable for clinical settings with limited resources. However, comparing it with the new o3 models is essential to determine the evolution of AI reasoning capabilities and their applicability in the pediatric field. Despite the extensive literature on the application of artificial intelligence in medicine, no analyses directly comparing the reasoning models o3-mini and o3-mini-high in handling pediatric clinical questions are currently available. This study, therefore, aims to fill this gap by providing a direct and in-depth comparison of the two models, with the objective of defining their potential contribution to therapeutic decision support in pediatric medicine.

## Materials and Methods

The study involved a total of 900 questions related to different pediatric clinical scenarios. The questions were drawn from the MedQA-USMLE dataset [5-6], covering a broad spectrum of conditions, from neonatal disorders to chronic diseases in childhood. The MedQA dataset was filtered to include only questions related to patients aged between 0 and 16 years. The total sample of 900 questions is divided into different categories, as shown in Figure 1.

**Figure 1.**
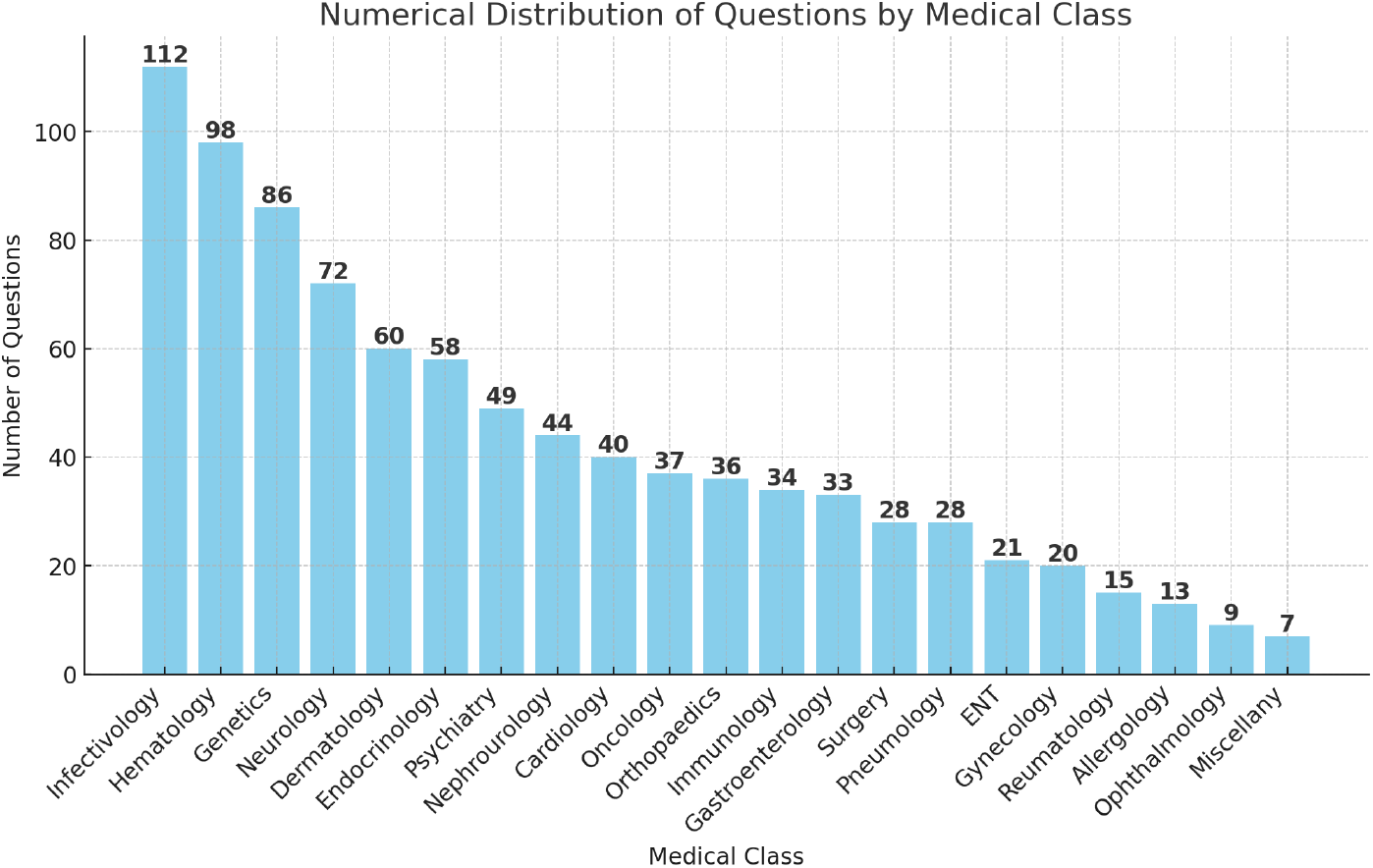
Categories of the 900 pediatric questions.

Through the evaluation of different categories, a detailed and balanced analysis of the performance of the o3-mini and o3-mini-high models was conducted in responding to various types of clinical questions. To ensure an adequate number of questions while keeping the number of weekly interactions within the limits imposed by the o3-mini-high model, the questions were presented in groups of 25. This strategy allowed the administration of the entire set of 900 questions in 36 sessions for each model, thus complying with the weekly usage restrictions. According to the information available at the time of writing, Plus users have access to 150 messages per day with o3-mini, while the limit for o3-mini-high is 50 messages per week [7]. This division enabled a precise evaluation of the models’ performance while keeping input parameters constant and limiting potential biases resulting from computational overload. o3-mini, designed to ensure an optimal balance between accuracy and latency, represents a lightweight yet effective solution for advanced reasoning tasks. On the other hand, o3-mini-high is a more advanced version, developed to provide a higher level of intelligence while maintaining optimized response times, making it particularly suitable for scenarios where accuracy is a priority. [2] The performance of the models was assessed using:

- **Accuracy:** This indicates the model’s ability to provide correct and relevant answers to clinical questions. This measure is generally expressed as a percentage, representing the ratio between correct responses and the total number of questions asked.
- **Latency:** This refers to the model’s response time, specifically the time elapsed between submitting the query and generating the response. The average response times for each model were measured using a digital stopwatch to ensure precision. In clinical settings, where timeliness can be crucial—such as in emergencies or triage—a low latency is essential to ensure that information is delivered quickly and effectively, contributing to timely clinical decisions.

The response analysis was conducted using a confusion matrix, which highlighted the agreements and discrepancies between the two models. The collected data were further subjected to a chi-square test (a statistical test to assess whether a significant difference exists between observed frequencies), including Yates’ correction, to verify the statistical significance of the observed differences. A p-value *<* 0.05 was considered significant.

## Results

The results of this study show that o3-mini-high achieves an accuracy of 90.55% (815 correct responses out of 900 questions), whereas o3-mini achieves an accuracy of 88.33% (795 correct responses out of 900 questions) (Figure 2 and Table 1). These findings indicate a higher accuracy for o3-mini-high.

**Table 1.** Comparisons of correct response rates among models.

| Models Comparisons | Correct Answers Model 1 | Correct Answers Model 2 | p-value |
| --- | --- | --- | --- |
| o3-mini (Model 1) vs o3-mini-high (Model 2) | 88,33% (795/900) | 90,55% (815/900) | p<0.00001 |

**Figure 2.**
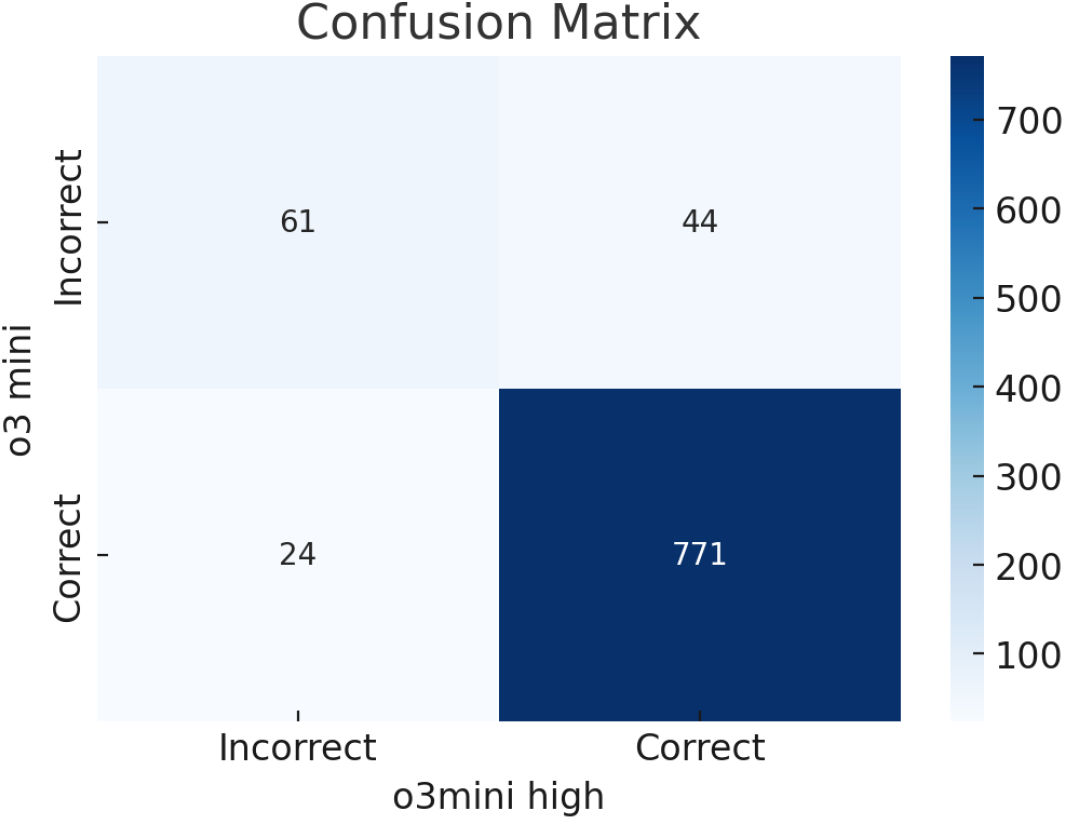
Accuracy comparison between o3-mini and o3-mini-high.

Figure 3 presents a radar graph comparing the distribution of correct responses per category between the two models, while Table 2 reports the number of correct responses for each question category in the analyzed models. Additionally, the response times of o3-mini-high were also shorter, with an average of 64.63 seconds compared to 71.63 seconds for o3-mini, marking a difference of approximately 7 seconds.

**Table 2.** Comparison between o3-mini-high and o3-mini in the distribution of correct responses per question category.

| Medical Class | o3-mini-high Correct/Total | o3-mini Correct/Total |
| --- | --- | --- |
| Allergology | 12/13 | 12/13 |
| Cardiology | 35/40 | 38/40 |
| Dermatology | 56/60 | 54/60 |
| ENT | 19/21 | 21/21 |
| Endocrinology | 52/58 | 52/58 |
| Gastroenterology | 25/33 | 24/33 |
| Genetics | 78/86 | 76/86 |
| Gynecology | 17/20 | 16/20 |
| Hematology | 86/98 | 88/98 |
| Immunology | 30/34 | 29/34 |
| Infectivology | 104/112 | 97/112 |
| Miscellany | 5/7 | 5/7 |
| Nephrourology | 44/44 | 42/44 |
| Neurology | 66/72 | 65/72 |
| Oncology | 35/37 | 30/37 |
| Ophthalmology | 9/9 | 8/9 |
| Orthopaedics | 34/36 | 34/36 |
| Pneumology | 26/28 | 24/28 |
| Psychiatry | 45/49 | 47/49 |
| Reumatology | 13/15 | 13/15 |
| Surgery | 24/28 | 20/28 |

**Figure 3.**
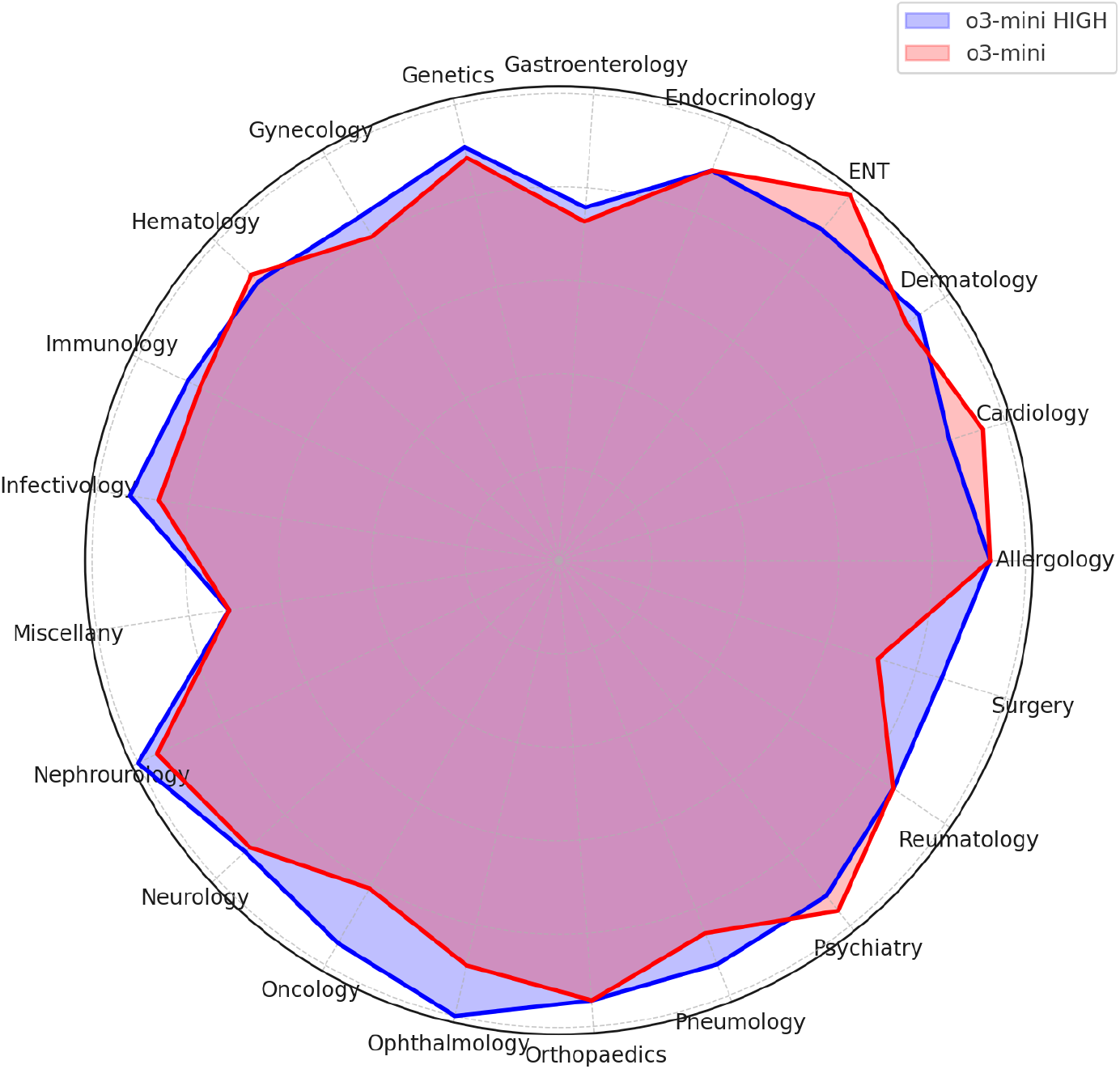
Radar graph comparing the distribution of correct responses between o3-mini-high (blue) and o3-mini (red) across different clinical categories. The graph highlights the performance differences between the two models.

The chi-square test yielded a value of 328.9675 with a p-value *<* 0.00001, confirming that the differences in performance between the two models are statistically significant. Even with Yates’ correction, the value remained significant at 322.5592 with a p-value *<* 0.00001. The discrepancy analysis showed that o3-mini-high corrected 44 errors made by o3-mini, while the latter corrected 24 errors introduced by the high version. The 61 shared errors represent a common critical area, suggesting potential limitations in the training data or in understanding complex pediatric scenarios.

## Discussion

The results of this study highlight how o3-mini-high not only offers greater overall accuracy, with 815 correct responses out of 900 pediatric questions, but also shorter response times, averaging 64.63 seconds compared to 71.63 seconds for o3-mini, with a 7-second reduction. This aspect is particularly relevant in pediatric clinical settings, where both precision and response speed are crucial. The confusion matrix analysis showed that, despite increased computational power and complexity, o3-mini-high improved time efficiency, providing a significant advantage for use in high-intensity clinical environments such as neonatal intensive care units and emergency departments, where every second saved can make a difference. o3-mini-high demonstrated the ability to maintain shorter response times while ensuring superior accuracy. The combination of speed and accuracy makes o3-mini-high the preferred choice in most pediatric clinical scenarios. The statistical analysis confirmed that the observed differences are not random, with a chi-square value of 328.9675 and a *p <* 0.00001), maintaining significance even after applying Yates’ correction. This statistical significance supports the idea that the algorithmic optimization of o3-mini-high not only improves accuracy but also enables more efficient processing, reducing response times. [6] This finding suggests that o3-mini-high could be an optimal option for clinical applications requiring both speed and reliability, such as pediatric triage, emergency management, and rapid consultations. Both models made 61 errors, of which 51 (83.6%) had the same incorrect response. Figure 4 illustrates the distribution of errors by clinical category.

**Figure 4.**
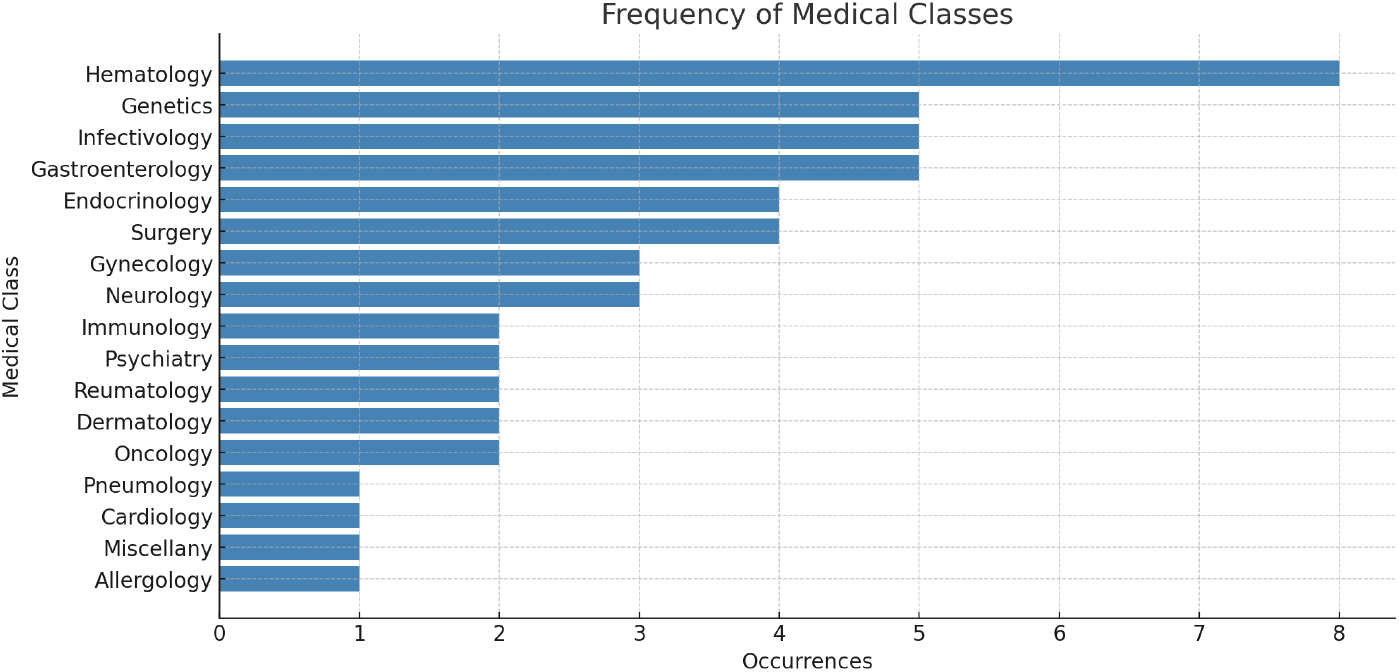
Distribution of errors by medical class. The chart shows the number of errors detected in each category, highlighting the specialties where the models encountered the greatest difficulties.

Additionally, it is important to consider the differences in accessibility and limitations between the analyzed models. The data related to DeepSeek-R1 comes from our previous study, in which the model demonstrated that its reasoning version is completely free, with no limits on the number of messages sent. This characteristic makes it particularly suitable for resource-limited settings or academic projects. [3,4] In contrast, OpenAI’s o3 models have some restrictions: o3-mini is available for free to all users, but Plus and Team users have a daily limit of 150 messages [7]. For o3-mini-high, usage limits may apply, although specific details are not clearly defined in the available sources. This difference in accessibility between DeepSeek-R1 and the o3 models could influence model selection based on the clinical context and available resources, an aspect that should be carefully considered during implementation. On the other hand, although o3-mini corrected some errors made by o3-mini-high (24 cases), it exhibited overall lower performance with longer response times. This could indicate that while o3-mini maintains an acceptable level of accuracy, it may not be the ideal choice in scenarios where speed is critical. However, the fact that both models made 61 common errors suggests that there are intrinsic limitations in the training data or the model architecture itself, which require further optimization. Future improvements should focus on enhancing the accuracy of o3-mini while maintaining its response efficiency and refining o3-mini-high to further reduce common errors. Additionally, implementing an ensemble system that combines the strengths of both models could provide an optimal solution, offering speed, accuracy, and reliability in complex clinical settings.

## Conclusions

This study compared the performance of o3-mini and o3-mini-high while integrating previous findings on DeepSeek-R1 to assess the effectiveness of reasoning models in managing pediatric clinical questions. o3-mini-high stood out for its superior accuracy and faster response times, making it an ideal choice for high-intensity settings such as pediatric emergencies. While o3-mini demonstrated lower performance, it maintains an acceptable level of accuracy and can be used in less critical scenarios. DeepSeek-R1, despite its slightly lower accuracy, remains a valuable option due to its accessibility and flexibility. Future implementation of ensemble systems could combine the strengths of each model, providing even more effective decision support, adaptable to the needs of modern pediatrics.

## Data Availability

All data produced in the present study are available upon reasonable request to the authors

